# Tocilizumab vs Bevacizumab in Critically Ill COVID-19 Patients: Registry-Based Prospective Study

**DOI:** 10.1101/2025.10.20.25338427

**Authors:** Yatendra Kumar Gupta, Raghubir Singh Khedar, Gulam Mateen Parihar, Anil Kumar Sharma, Pramod Sarwa, Rajeev Gupta

**Affiliations:** Department of Critical Care, and Eternal Hospital, Eternal Heart Care Centre & Research Institute, Jaipur, India; Department of Medicine, Eternal Hospital, Eternal Heart Care Centre & Research Institute, Jaipur, India

**Author notes:** Correspondence:* Dr Rajeev Gupta, Department of Medicine, M-Floor, Eternal Heart Care Centre & Research Institute, Jagatpura Road, Jawahar Circle, Jaipur 302017 India.

**Keywords:** SARS-CoV-2, COVID-19, coronavirus, tocilizumab, bevacizumab, cytokine storm

## Abstract

**Objectives:** Tocilizumab, an interleukin-6 (IL-6) inhibitor, and bevacizumab, a vascular endothelium growth factor (VEGF) inhibitor, have been used in critically ill COVID-19 patients for cytokine storm. We performed a registry-based prospective study to compare the efficacy of these two drugs.

**Methods:** Virologically confirmed hospitalised patients with severe COVID-19 who received either tocilizumab or bevacizumab have been included. Management details and outcomes were recorded. The primary outcome was in-hospital deaths and secondary outcomes were 30-day deaths, changes in oxygen requirement at 48 hours of drug administration, and duration of ICU stay. Descriptive statistics are reported.

**Results:** 1345 COVID-19 patients were hospitalised during the study period, 87 with severe COVID-19 received tocilizumab(n=62) or bevacizumab(n=25). Patients in the tocilizumab group were older (62.6+11vs.53.2+14, p=0.001) with more cardiovascular disease and no significant differences in clinical features, laboratory investigations, or radiological and biochemical markers of disease severity. Oxygenation, ventilatory support and proning were similar as were supportive therapies (steroids, remdesivir, anticoagulants)(p>0.05). In-hospital deaths in tocilizumab vs bevacizumab group were 47(75.8%) vs 12(48.0%)(p=0.012) with unadjusted hazard ratio(HR) 1.91 (95% CI 0.99-3.65, p=0.050) and age-adjusted HR 1.76 (0.90-3.44, p=0.097). Secondary outcome of 30-day death was also more in tocilizumab group: 49(79.0%) vs 13(52.0%) (p=0.011). Within-group comparison showed that PaO_2_:FiO_2_ at 48-hour of drug administration was better in bevacizumab group (p=0.003) compared to tocilizumab(p=0.651).

**Conclusion:** VEGF inhibitor bevacizumab led to significantly better ventilatory outcomes and lower in-hospital and 30-day deaths compared to IL-6 inhibitor tocilizumab in severely ill COVID-19 patients. Randomised studies are required to confirm these findings.

## INTRODUCTION

A substantial proportion of patients with COVID-19 develop acute respiratory distress syndrome (ARDS) leading to hypoxia, organ dysfunction and death.^1,2,3,4,5^ Severe COVID-19 is associated with clinical symptoms of ARDS, radiological abnormalities and raised biomarkers-neutrophil-lymphocyte ratio, hs-CRP (highly sensitive C reactive protein), d-dimer, interleukin-6 (IL-6), lactate dehydrogenase (LDH), ferritin, etc.^4,5,6,7,8^ Various treatments have been evaluated to combat the high death rate associated with severe COVID-19. The management strategies have focused on pathophysiological pathways of severe disease such as microvascular thrombosis, inflammatory cytokine storm, and vascular permeability.^10,11^ Large clinical trials and meta-analyses have supported use of dexamethasone,^12^ anticoagulants,^13^ and non-invasive oxygenation therapies, including proning, in severe COVID-19.^10,11^ Most of the other pharmaceutical-based strategies have provided equivocal results.^10,11^ Tocilizumab, an IL-6 inhibitor, has been proposed as a treatment option due to its ability to modulate the inflammatory cytokine response.^14^ However, randomized trials have shown mixed results regarding its effectiveness in improving clinical outcomes,^15-25^ although the RECOVERY trial reported mortality benefit with tocilizumab in critically ill COVID-19.^24^

Bevacizumab an anti-VEGF (vascular endothelial growth factor) drug and has been evaluated in severe adult respiratory distress syndrome.^26-28^ Only a few small trials regarding the efficacy of bevacizumab in COVID-19 have been reported with inconclusive outcomes.^29-32^ We performed a registry-based real-world comparative evaluation of tocilizumab and bevacizumab in severe COVID-19. The primary objective of the study was to compare the efficacy of tocilizumab vs bevacizumab on in-hospital deaths. Secondary objectives were overall deaths at 30 days post-discharge, effect on oxygenation status at the end of 48 hours of drug administration and duration of stay in the intensive care unit.

## METHODS

This is a single-centre, real-world, evidence-based observational study conducted in India. We initiated a registry of all hospitalised patients with COVID-19 since the beginning of the epidemic in India (April 2020).^5^ The protocol was approved by the institutional ethics committee (Government of India registration, CDSCO No. ECR/615/Inst/RJ/2014/RR-20). Informed consent from the patient or next of kin was obtained. We specifically obtained permission for the publication of anonymized data as permitted by the ethics committee. The registry data have been published previously.^5,9^

For the present study, we included patients aged >18 years with virologically confirmed COVID-19.^9^ Major inclusion criteria were patients admitted to medical intensive care units (ICU), high-resolution computed tomography (HRCT) detected lung infiltrates of more than 50%, PaO_2_:FiO_2_ ratio of 50-300 mm Hg, and all those patients who received either tocilizumab or bevacizumab during the hospital stay. Decision to use tocilizumab or bevacizumab was at the discretion of the treating clinician depending on availability and affordability of the drugs. The drugs were administered after obtaining informed consent from patient’s attendants and exclusion of contraindications. Exclusion criteria were refusal of consent by patients (or relatives), suspected allergy to tocilizumab or bevacizumab, patients with any malignancy, severe liver disease (transaminase levels >5 times upper limit of normal or raised Child-Pugh score), acute or chronic renal disease needing dialysis, active bleeding, active gastric ulcers, extrapulmonary source of sepsis, active tuberculosis, congestive heart failure, myocardial infarction within one year, active hepatitis or HIV infection, and pregnant or lactating mothers. Severe COVID-19 was defined as patients with SpO_2_ <90% and requiring oxygen support. Patients in both groups received similar standards of care and treatment protocols as reported previously.^5^ Besides routine laboratory investigations, markers of inflammation including neutrophil-lymphocyte ratio, hsCRP (high sensitivity C-reactive protein), d-dimer, IL-6, ferritin, lactate dehydrogenase were measured.^9^ HRCT scans of thorax were performed in all the patients.

The treatment included optimum hydration, intravenous steroids (dexamethasone 6 mg/day or equivalent doses of other steroids), intravenous remdesivir (200 mg on day 1, followed by 100 mg daily for 5 days),^34^ anticoagulation (unfractionated heparin or enoxaparin)^13^ unless contraindicated. Respiratory support was provided according to the clinical condition with or without proning and included non-invasive ventilation, high flow nasal cannula oxygenation, or invasive mechanical ventilation. Awake proning was done in non-ventilated patients. Non-evidence based therapies (hydroxychloroquine, ivermectin or plasma therapy) were not used.^5^ Tocilizumab was administered as a single intravenous dose over 60 minutes as infusion. The dose was calculated according to the body weight: 800 mg for >90 kg; 600 mg for ≤90 to 65 kg; 400 mg for ≤65 to >40 kg; and 8 mg/kg for weight ≤40 kg. Bevacizumab was administered intravenously at 7.5 mg/kg.

Apart from enumeration of in-hospital deaths and 30-day follow-up deaths, we also determined the ratio of PaO_2_ (arterial partial pressure of oxygen) to FiO_2_ (fraction of inspired oxygen) before the drug was administered and at the end of 48 hours of the drug administration. If the patient expired during this period, PaO_2_:FiO_2_ ratio just before the death was recorded. We also noted complications related to these drugs. The mortality data at 30-day of discharge was obtained by telephonic call, and were available for all except 2 in the tocilizumab group (response rate 97.7%).

### Statistical analysis

All data were entered into MS-Excel worksheets and transferred to SPSS statistical program (version 22.0) for further analyses. Demographic and clinical characteristics were reported using means and standard deviations (SD) for normally distributed continuous variables, medians and 25-75 interquartile range (IQR) for skewed variables, numbers and per cent for categorical variables.^9^ Data were analyzed using t-test for continuous variables with normal-distribution and Kruskal-Wallis test or continuous variables with skewed distribution. χ^2^ test was performed for identification of significance of inter-group differences in categorical variables. Wilcoxon signed ranked test for used for within-group before-after comparisons. Hazard ratios (HR) and 95% confidence intervals (CI) for outcomes were calculated using Cox regression to compare the primary outcome in tocilizumab vs bevacizumab groups. We performed step-wise hazard ratio calculations before adjustment (unadjusted), age-adjusted, age-sex adjusted, age-sex and CVD adjusted and multivariate-adjusted (age, sex, risk factors, comorbidities and treatment). P value <0.05 is considered significant.

## RESULTS

During the 28-month study period (April 2020 to July 2022) 1345 virologically confirmed COVID-19 patients were hospitalized. Of these, 87 patients received either tocilizumab (n=62) or bevacizumab (n=25) and have been included in the study (Figure 1). The baseline demographic details, clinical status and laboratory investigations are in Table 1. Patients in the tocilizumab group were older (62.6+11vs.53.2+14, p=0.001) with more patients >60 years (64.5 vs 32.0%, p=0.006). There were more patients with cardiovascular diseases (CVD) in the tocilizumab group (n=16, 25.8%) compared to bevacizumab group (n=1, 4.0%) (p=0.020). No significant differences were observed in prevalence of other co-existing diseases, clinical features and laboratory investigations. Haematological and biochemical markers of severe disease-raised neutrophil-lymphocyte ratio and raised levels of hsCRP, d-dimer, interleukin-6, ferritin and lactic dehydrogenase-are also not significantly different. HRCT scan score (out of 25) were also similar in the two groups.

**Table 1:**
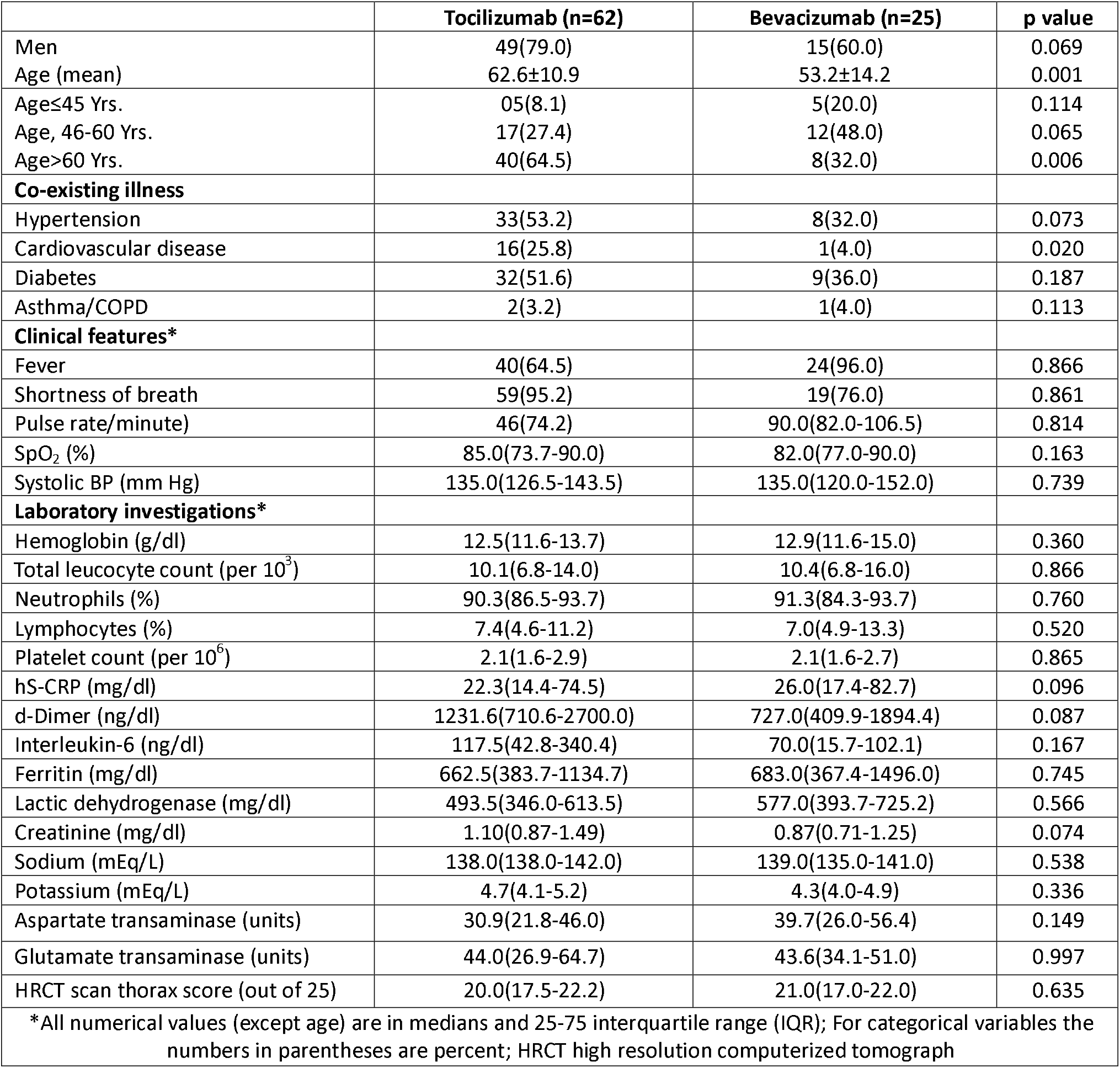
Baseline demographic, clinical and laboratory variables in tocilizumab and bevacizumab groups.

|  | **Tocilizumab (n=62)** | **Bevacizumab (n=25)** | **p value** |
| --- | --- | --- | --- |
| Men | 49(79.0) | 15(60.0) | 0.069 |
| Age (mean) | 62.6±10.9 | 53.2±14.2 | 0.001 |
| Age≤45 Yrs. | 05(8.1) | 5(20.0) | 0.114 |
| Age, 46-60 Yrs. | 17(27.4) | 12(48.0) | 0.065 |
| Age>60 Yrs. | 40(64.5) | 8(32.0) | 0.006 |
| **Co-existing illness** |  |  |  |
| Hypertension | 33(53.2) | 8(32.0) | 0.073 |
| Cardiovascular disease | 16(25.8) | 1(4.0) | 0.020 |
| Diabetes | 32(51.6) | 9(36.0) | 0.187 |
| Asthma/COPD | 2(3.2) | 1(4.0) | 0.113 |
| **Clinical features\*** |  |  |  |
| Fever | 40(64.5) | 24(96.0) | 0.866 |
| Shortness of breath | 59(95.2) | 19(76.0) | 0.861 |
| Pulse rate/minute) | 46(74.2) | 90.0(82.0-106.5) | 0.814 |
| SpO<sub>2</sub> (%) | 85.0(73.7-90.0) | 82.0(77.0-90.0) | 0.163 |
| Systolic BP (mm Hg) | 135.0(126.5-143.5) | 135.0(120.0-152.0) | 0.739 |
| **Laboratory investigations\*** |  |  |  |
| Hemoglobin (g/dl) | 12.5(11.6-13.7) | 12.9(11.6-15.0) | 0.360 |
| Total leucocyte count (per 10<sup>3</sup>) | 10.1(6.8-14.0) | 10.4(6.8-16.0) | 0.866 |
| Neutrophils (%) | 90.3(86.5-93.7) | 91.3(84.3-93.7) | 0.760 |
| Lymphocytes (%) | 7.4(4.6-11.2) | 7.0(4.9-13.3) | 0.520 |
| Platelet count (per 10<sup>6</sup>) | 2.1(1.6-2.9) | 2.1(1.6-2.7) | 0.865 |
| hS-CRP (mg/dl) | 22.3(14.4-74.5) | 26.0(17.4-82.7) | 0.096 |
| d-Dimer (ng/dl) | 1231.6(710.6-2700.0) | 727.0(409.9-1894.4) | 0.087 |
| Interleukin-6 (ng/dl) | 117.5(42.8-340.4) | 70.0(15.7-102.1) | 0.167 |
| Ferritin (mg/dl) | 662.5(383.7-1134.7) | 683.0(367.4-1496.0) | 0.745 |
| Lactic dehydrogenase (mg/dl) | 493.5(346.0-613.5) | 577.0(393.7-725.2) | 0.566 |
| Creatinine (mg/dl) | 1.10(0.87-1.49) | 0.87(0.71-1.25) | 0.074 |
| Sodium (mEq/L) | 138.0(138.0-142.0) | 139.0(135.0-141.0) | 0.538 |
| Potassium (mEq/L) | 4.7(4.1-5.2) | 4.3(4.0-4.9) | 0.336 |
| Aspartate transaminase (units) | 30.9(21.8-46.0) | 39.7(26.0-56.4) | 0.149 |
| Glutamate transaminase (units) | 44.0(26.9-64.7) | 43.6(34.1-51.0) | 0.997 |
| HRCT scan thorax score (out of 25) | 20.0(17.5-22.2) | 21.0(17.0-22.0) | 0.635 |
| *All numerical values (except age) are in medians and 25-75 interquartile range (IQR); For categorical variables the numbers in parentheses are percent; HRCT high resolution computerized tomograph |  |  |  |

**Figure 1.**
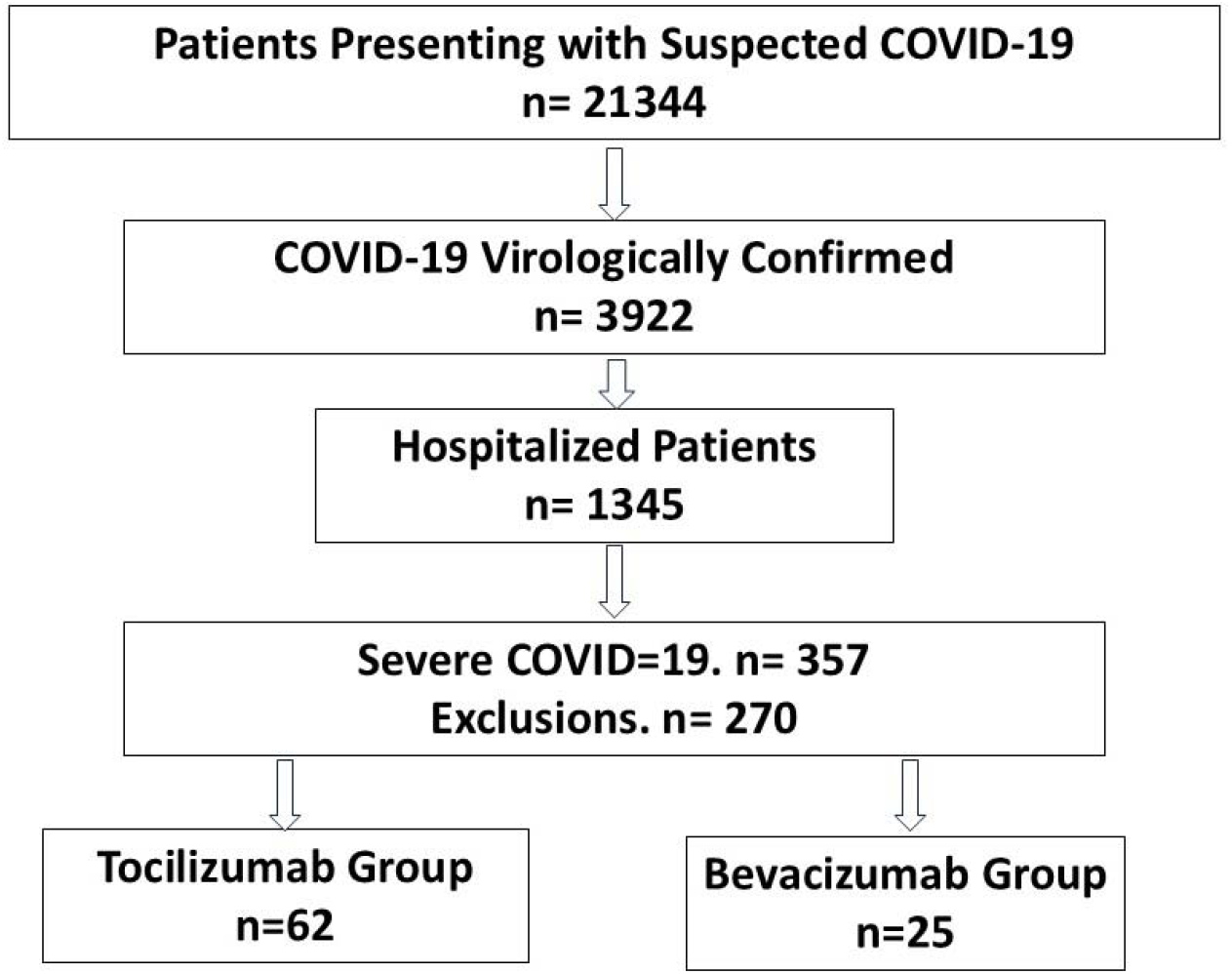
CONSORT flow-diagram of the study participants.

Details of clinical management are in Table 2. All the patients were on at least one of three oxygenation support strategies-high flow nasal cannula, non-invasive ventilation or invasive mechanical ventilation. There was no significant difference in the type of respiratory support in the treatment groups (Table 2). In tocilizumab vs bevacizumab groups, before drug administration, median duration of oxygen support was 6.0 (5.0-10.0) vs 6.0 (4.0-9.0) days; median oxygen saturation (SpO_2_) was 85 (73.7-90%) vs 82 (77-90%), and median PaO_2_ was 90.0 (77.2-104.5) vs 96.0 (72.0-118.0) (p=n.s., Table 2). All the study participants received steroids and anticoagulants. Intravenous remdesivir was administered to 60 of 62 patients in the tocilizumab group and in all in the bevacizumab group. Most patients in both groups were provided proning for improvement of oxygenation. No difference on adverse outcomes was noted.

**Table 2:**
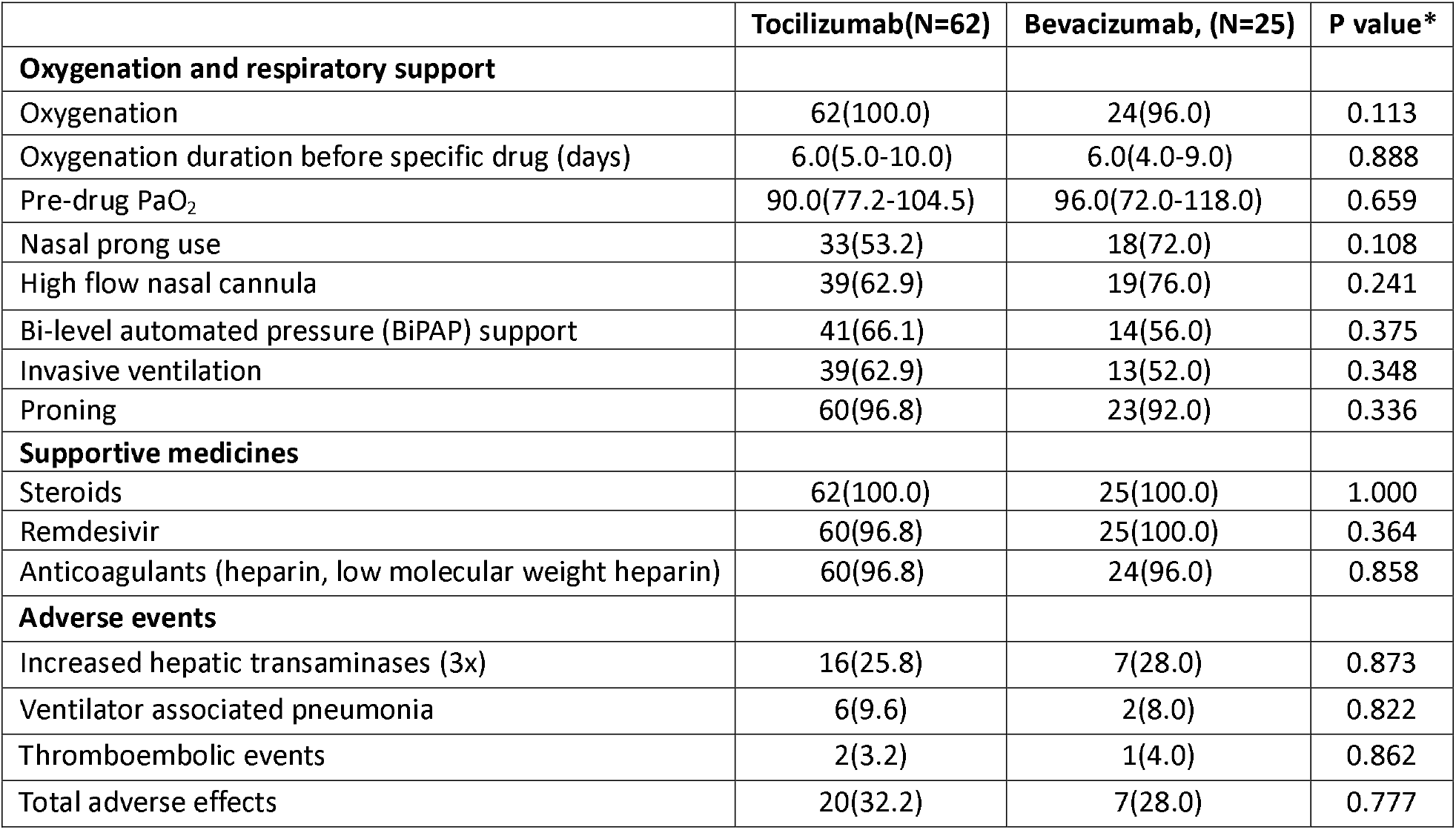
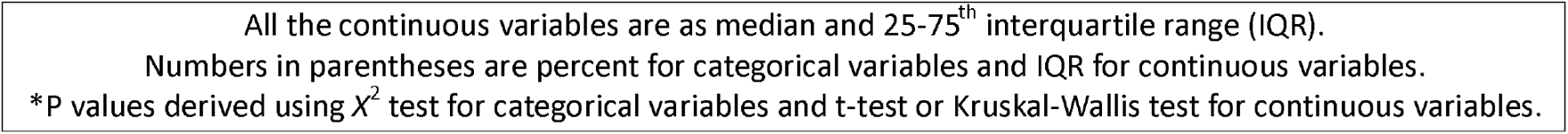
Clinical features and management in the two groups.

|  | **Tocilizumab(N=62)** | **Bevacizumab, (N=25)** | **P value\*** |
| --- | --- | --- | --- |
| **Oxygenation and respiratory support** |  |  |  |
| Oxygenation | 62(100.0) | 24(96.0) | 0.113 |
| Oxygenation duration before specific drug (days) | 6.0(5.0-10.0) | 6.0(4.0-9.0) | 0.888 |
| Pre-drug PaO<sub>2</sub> | 90.0(77.2-104.5) | 96.0(72.0-118.0) | 0.659 |
| Nasal prong use | 33(53.2) | 18(72.0) | 0.108 |
| High flow nasal cannula | 39(62.9) | 19(76.0) | 0.241 |
| Bi-level automated pressure (BiPAP) support | 41(66.1) | 14(56.0) | 0.375 |
| Invasive ventilation | 39(62.9) | 13(52.0) | 0.348 |
| Proning | 60(96.8) | 23(92.0) | 0.336 |
| **Supportive medicines** |  |  |  |
| Steroids | 62(100.0) | 25(100.0) | 1.000 |
| Remdesivir | 60(96.8) | 25(100.0) | 0.364 |
| Anticoagulants (heparin, low molecular weight heparin) | 60(96.8) | 24(96.0) | 0.858 |
| **Adverse events** |  |  |  |
| Increased hepatic transaminases (3x) | 16(25.8) | 7(28.0) | 0.873 |
| Ventilator associated pneumonia | 6(9.6) | 2(8.0) | 0.822 |
| Thromboembolic events | 2(3.2) | 1(4.0) | 0.862 |
| Total adverse effects | 20(32.2) | 7(28.0) | 0.777 |

All the continuous variables are as median and 25-75<sup>th</sup> interquartile range (IQR).
Numbers in parentheses are percent for categorical variables and IQR for continuous variables.
\*P values derived using $\chi^2$ test for categorical variables and t-test or Kruskal-Wallis test for continuous variables.

A comparison of clinical outcomes in the two groups is shown in Table 3. The primary outcome of in-hospital mortality in tocilizumab group was 47 (75.8%) vs 12 (48.0%) in bevacizumab group (p=0.012). The unadjusted hazard ratio for in-hospital deaths in tocilizumab vs bevacizumab was 1.91 (95% CI 0.99-3.65, p=0.050). Stepwise analyses show that although the HR declined with adjustment for age (HR 1.76, 0.90-3.44, p=0.097), age and sex (HR 1.53, 0.76-3.10, p=0.234), age-sex and CVD (HR 1.41, 0.69-2.89), p=0.347), and multiple factors (age, sex, risk factors and co-morbidities) (HR 1.21, 0.58-2.53, p=0.609), it remained substantial (Figure 2). The secondary outcome of overall deaths at 30 days post-discharge was also significantly more in the tocilizumab group 49 (79.0%) compared to bevacizumab (13 (52.0%) (p=0.011). Between-group comparison of the PaO_2_:FiO_2_ ratio at 48-hour after tocilizumab and bevacizumab drug administration in the two groups were not statistically significant (median 83.0, IQR 71-0-114.0 vs 138.0, IQR 73-178.0, p=0.360). However, within-group comparisons show that bevacizumab led to better improvement (Wilcoxon signed-rank test, p=0.003) compared to tocilizumab (p=0.651). No significant differences were noted in the duration of hospitalization as well as duration of ICU stay and in adverse events such as secondary infections and thromboembolic complications.

**Table 3:** Study outcomes in tocilizumab and bevacizumab groups.

|  | Tocilizumab(N=62) | Bevacizumab, (N=25) | P value* |
| --- | --- | --- | --- |
| **Primary outcomes** |  |  |  |
| In-hospital deaths | 47(75.8) | 12(48.0) | 0.012 |
| **Secondary outcomes** |  |  |  |
| 30-day overall deaths | 49(79.0) | 13(52.0) | 0.011 |
| 48 hr post-drug PaO<sub>2</sub>: FiO<sub>2</sub> ratio | 83.0(71.0-114.0) | 138.0(73.0-178.0) | 0.360 |
| Pre-drug and post-drug PaO<sub>2</sub>: FiO<sub>2</sub> comparison** | 0.45 (p=0.651) | 3.0 (p=0.003) | -- |
| Duration of hospitalization | 14.0(9.0-18.0) | 15.0(11.0-22.0) | 0.760 |
| Duration of ICU stay | 13.0(8.0-17.0) | 13.0(9.0-17.5) | 0.434 |
| All the continuous variables are in median and 25-75<sup>th</sup> interquartile range (IQR).<br>Numbers in parentheses are percent for categorical variables and IQR for continuous variables.<br>*P values derived using $\chi^2$ test for categorical variables and Kruskal-Wallis test for continuous variables;<br>**Before-after comparison for each drug performed using Wilcoxon sign-rank test; |  |  |  |

**Figure 2.**
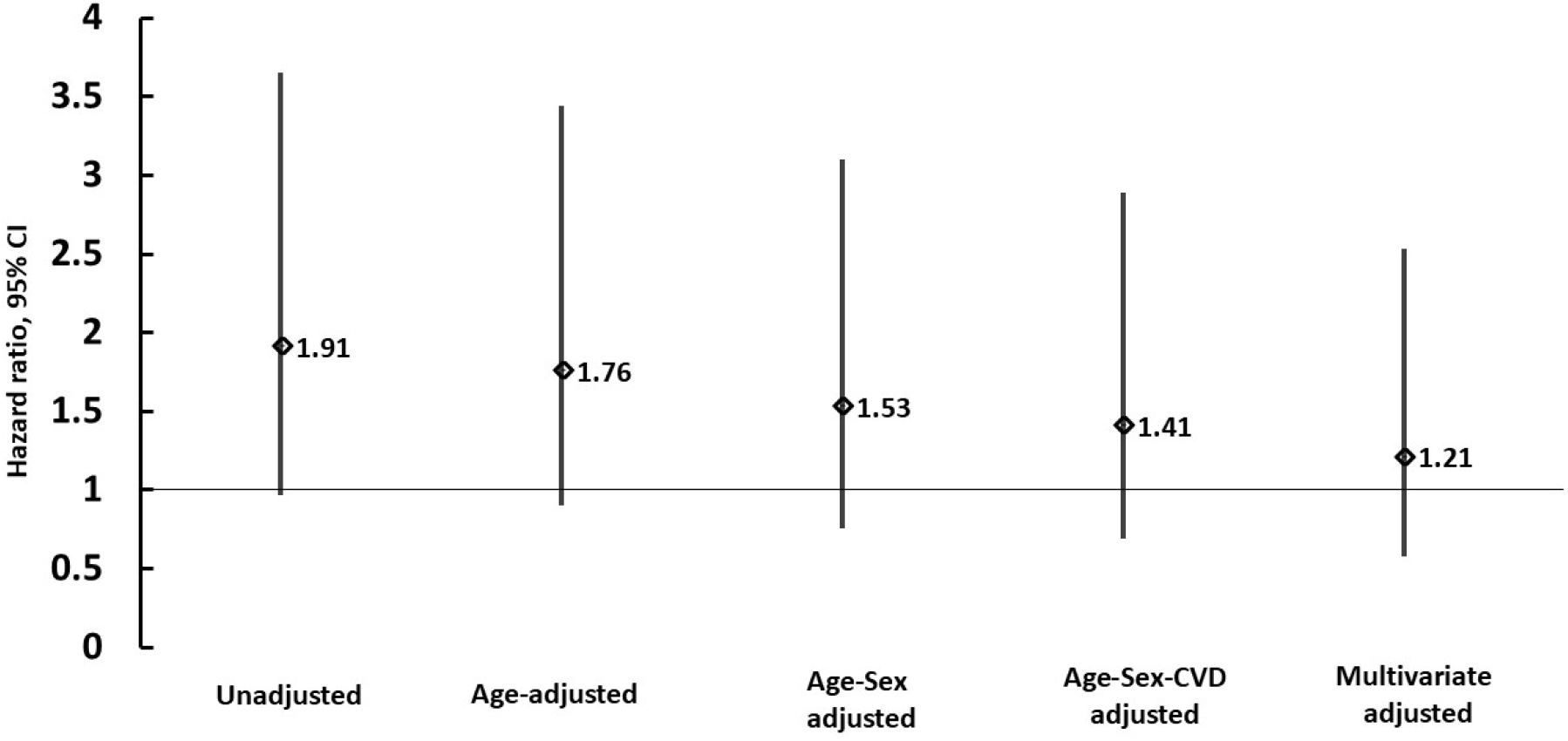
Hazard ratio of in-hospital deaths in tocilizumab vs bevacizumab groups (Cox regression analyses, univariate and adjusted outcomes). Multivariate adjustment includes age, sex, cardiovascular diseases (CVD), risk factors and comorbidities

## DISCUSSION

This study shows that treatment with bevacizumab is better than tocilizumab for the reduction of in-hospital and 30-day deaths in critically ill COVID-19 patients. The improvement in PaO_2_:FiO_2_ ratio following bevacizumab also correlates with its mortality benefit. All patients in the study had haematological, biochemical and radiological markers of severe COVID-19 and the pre-therapy mean PaO_2_ :FiO_2_ ratio was low confirming severe ARDS.^34^

Previous non-randomised studies provided initial evidence that tocilizumab reduced progression to invasive mechanical ventilation or death and shortened time to discharge,^23^ although other studies reported mixed results.^15-22^ This was attributed to variations in the dose of tocilizumab, stage of disease in which the drug was administrated, and differences in the severity of COVID-19. Patients who were administered tocilizumab in the late stages of the disease with multiorgan dysfunction had poor outcomes.^20,21^ In the RECOVERY trial tocilizumab reduced 28-day mortality in patients on oxygen support.^24^ Conversely, some other randomized trials have not shown benefit of tocilizumab on primary outcomes,^15,23^ and in a randomized trial in Brazil^25^ mortality at 15 days was increased in the group assigned to tocilizumab. In the STOP-COVID trial tocilizumab was associated with decreased mortality only among patients when the duration of onset of symptoms to ICU admission was less than three days.^20^ No benefit was observed in randomized trials when the duration of symptoms onset to therapy was more than 8 days.^21,35,36^ In the present study, both tocilizumab and bevacizumab were administered at a mean of 6 days following admission. This was due to waiting for the development of hypoxia as many of moderate to severely ill COVID-19 patients improved with standard care. Previous studies have also compared tocilizumab administration at different COVID-19 severity levels. In the COVACTA trial,^35^ patients with a wide spectrum of illness were included ranging from those with moderate hypoxia to invasive mechanical ventilation and reported uncertain benefit of tocilizumab. On the other hand, in the EMPACTA trial,^23^ enrolled patients were not receiving mechanical ventilation at baseline and led to better outcomes. In our study, tocilizumab was administered to patients with severe hypoxia with mean P:F ratio of 92.0 (IQR 75.0-106.0). In India, as in the present study, uncertainty about the efficacy of tocilizumab, high cost, and limited availability are important factors to restrict early use of tocilizumab.^37,38^

Only limited and small studies have been conducted for bevacizumab in severe COVID-19. This could be due to its availability in the later stages of the epidemic and off-label use in South Asian countries only. A non-randomised study from India,^30^ did not find bevacizumab superior to standard care in reducing mortality although radiological improvement and an improvement in P:F ratio was observed.^30^ In a comparative observational study in Bangladesh, outcomes were better following bevacizumab compared to tocilizumab.^31^ VEGF promotes the expression of endothelial cell anti-apoptotic proteins and blocks apoptosis of endothelial cells which may be useful in COVID-19. VEGF-genetic polymorphisms contribute to the development of acute respiratory distress syndrome (ARDS) and severe hypoxia. Studies showed that the presence of genetic polymorphism is associated with a high risk of pulmonary complications, including ARDS.^26,27,28^ In the present study, although the two groups were different in terms of age, but were similar in terms of oxygenation status and degree of lung involvement before drug administration and had identical levels of biochemical markers of severity (Table 2) and shows baseline comparability of the two groups.

The study has limitations other than those mentioned above. This is a single-centre observational study and not a randomized comparison. Patients who received bevacizumab were younger although age-adjusted comparisons show superiority of the molecule (although not significant (Figure 2). Patients with preexisting cardiovascular disorders were more in the tocilizumab group (Table 1) and this might have contributed to its higher mortality. Tocilizumab and bevacizumab were not given to all critically ill patients due to either non-availability or affordability.^37^ There were no defined criteria for the choice of either of the drugs in our patients, similar to Indian multi-site registries,^38^ and this is also a limitation. A small sample size in both groups is the main limitation of our study. On the other hand, our study suggests that VEGF signalling pathways should be further explored in various acute respiratory distress syndrome (ARDS) stages to better understand VEGF regulation and the development of novel anti-VEGF therapies beyond bevacizumab. A larger study on this anti-VEGF drug before recommending it strongly as a treatment option. Our study suggests the need for well-designed randomised controlled trials for the evaluation of VEGF inhibitors in ARDS patients in general and severe COVID-19 in particular.

## Data Availability

All data produced in the present work are contained in the manuscript

